# Opioid crisis in Germany? Insights from a cross-sectional nationwide survey within the German drug scene

**DOI:** 10.64898/2026.05.28.26354329

**Authors:** Jeanette Röhrig, Lorenz Sutter, Nicolas Witsch, Lena Rademacher, Maurice Cabanis

## Abstract

**Introduction:** Synthetic opioids cause tens of thousands of deaths annually in North America and appear to be increasing in Europe. This study examined high-risk substance use in the German drug-using community with a particular focus on the synthetic opioids fentanyl and nitazenes.

**Methods:** A cross-sectional online survey was conducted in the drug-using community in Germany (August–September 2025). The sample included 235 individuals aged ≥14 years (mean age=43.4 years; 57.9% male), recruited by peers in open drug-use scenes (79.6%) and in addiction clinics and opioid substitution services. The primary outcome was past 12-month substance use. Additional measures included opioid sources, forms, routes of administration, perceived changes in availability and price, risk perceptions, overdose experiences, and harm-reduction behavior.

**Results:** 227 respondents reported substance use (mean: 6.2 substances). 73.1% (95% CI=67.0– 78.5%) had used at least one opioid in the past year. Synthetic opioids were reported across regions and all age and gender groups. Among participants reporting a shortage of their primary opioid, 25.0% (95% CI=15.8–37.2%) used fentanyl as a substitute. 56.5% (95% CI=36.8–74.3%) of synthetic opioid users reported an overdose in the past 12 months. Most respondents perceived synthetic opioids as highly risky and feared contamination, but only 9.9% (95% CI=6.6–14.7%) use drug-checking services.

**Conclusions:** Synthetic opioids have entered the German drug scene and are associated with high overdose rates and limited access to harm-reduction measures. These findings indicate an emerging synthetic opioid transition in Germany and highlight the need for expanded surveillance, naloxone distribution, and drug-checking services.

## INTRODUCTION

Opioids account for a substantial share of global drug-related mortality, with approximately two-thirds of more than 500,000 annual drug-related deaths linked to opioid use [1]. Synthetic opioids pose particularly high risks due to their extreme potency: fentanyl is estimated to be 50–100 times more potent than morphine [2], and several nitazene derivatives exceed even this level [3,4]. The risks are amplified by the frequent adulteration of illicit drug supplies, where people who use drugs (PWUD) are often unaware of the presence, concentration, or identity of synthetic opioids in their substances [5,6].

The North American overdose crisis, driven increasingly by illicitly manufactured fentanyl and its analogues, continues to cause tens of thousands of deaths annually despite recent declines in the United States [7,8]. Whether a similar trajectory could unfold in Europe has become a pressing public health question. In Europe, heroin remains the most commonly used illicit opioid and the primary driver of opioid-related health harms [9]. However, monitoring data indicate a progressively more diverse opioid market, with increasing misuse of opioid medications and the continuous emergence of highly potent synthetic opioids, including fentanyl analogues and nitazenes. Because synthetic opioids can be manufactured in small laboratories using widely available precursors and transported in minimal quantities, even small suppliers can rapidly affect regional markets [10].

In Germany, 2,137 drug-related deaths were recorded in 2024, the second highest number after the historic peak of 2,227 deaths in 2023. Of these, 154 were attributable to new psychoactive substances, representing an approximately 70% increase over the previous year, with 36% of these deaths linked to fentanyl [10]. European monitoring data identify Germany as having the highest number of fentanyl-related deaths among EU Member States reporting to the European Union Drugs Agency [9]. In parallel, drug-related deaths among people under 30 years of age are rising, with synthetic opioids and new psychoactive substances playing an increasing role in toxicological findings [11]. The number of clandestine drug laboratories detected by police in Germany also increased from 14 in 2023 to 25 in 2024 [10].

These developments indicate that the synthetic opioid landscape in Germany is changing, but existing routine statistics are insufficient to capture rapid shifts in substances, use patterns, and regional dynamics [11,12]. Scene-based surveys that reach PWUD remain underutilized in Germany; to date, such surveys have been conducted only in individual cities [13,14]. This nationwide survey examined high-risk substance use in Germany’s drug scenes, with a focus on the prevalence and geographic distribution of synthetic opioid use, particularly fentanyl and nitazenes, perceived changes in availability, overdose experiences, risk perceptions, and use of harm-reduction services.

## METHODS

### Study Design

We conducted an exploratory online survey without confirmatory hypotheses. The design was cross-sectional and observational, with decentralized recruitment across multiple locations in Germany. Recruitment settings comprised open drug-use scenes and medical care settings. The study was conducted in cooperation with large national and local peer and family advocacy organizations which established contact with peer interviewers. These were familiar with local drug-use scenes and recruited in their communities. All peer interviewers were offered standardized training prior to data collection, covering the study objectives, questionnaire content, ethical considerations, and practical aspects of recruitment and survey administration. In addition, the three largest addiction treatment clinics offering a qualified detoxification treatment in each federal state and all diamorphine-prescribing practices across Germany (n=17) were contacted and asked to hang up posters and distribute information cards with a link to the online survey. Questionnaire administration and data management were centrally coordinated by the study team. Participation was voluntary, and informed consent was obtained via an explicit opt-in step before accessing the questionnaire. The study protocol was approved by the Ethics Committee of the State Medical Association (Landesärztekammer) Baden-Württemberg (approval number: F-2024-120).

### Participants

Eligible participants were aged 14 years or older and currently connected to drug-use scenes through use, treatment, or harm-reduction services. Eligibility was determined by trained peer interviewers during recruitment or self-report within the online questionnaire. Geolocation was recorded automatically when enabled; otherwise, participants entered their location manually.

### Data Collection

Data were collected using a structured online questionnaire in SoSci Survey [15]. The questionnaire was accessible via QR code or direct link and could be completed on participants’ own devices or on devices provided by peer interviewers. Data collection was conducted between 4th August 2025 and 21st September 2025.

### Outcome Measures

The primary outcome was self-reported substance use in the past 12 months, assessed by a multiple-choice item. Secondary outcomes comprised indicators related to (synthetic) opioid use, associated risk perceptions and behavior, and overdose-related experiences. Participants who reported the use of fentanyl/carfentanyl, nitazenes and/or another opioid were asked about their primary opioid and alternative substances used when the primary substance was unavailable. Additional items assessed sources and forms of opioids, routes of administration, and perceived changes in availability and price. Among those who have not consumed synthetic opioids, self-reported interest in fentanyl and nitazenes consumption, awareness and discussion of fentanyl within peer networks, perceived availability and fears regarding adulteration in other drugs were assessed. Finally, experiences with overdose (personal or within the social environment), and general risk consumption behavior, overdose-related concerns and suicide attempts were investigated.

### Questionnaire Development

The questionnaire was developed through an iterative expert-consensus process, involving physicians, psychotherapists, social workers, researchers, people with lived experience of substance use, and representatives of affected families. The instrument was reviewed in two consensus meetings and pretested in cognitive walkthroughs with two members of the target population. Feedback was used to refine wording, response options, and survey flow.

### Data analysis

Statistical analyses were conducted in SPSS version 27.0 and were descriptive. Categorical variables are reported as percentages with 95% Wilson confidence intervals based on valid responses; continuous variables are reported as means with standard deviations. Descriptive subgroup analyses were calculated for age and gender groups.

## RESULTS

### Participants

A total of 260 individuals accessed the survey. Of these, 25 (9.6%) did not provide any further information besides demographics and were excluded, yielding a final analytic sample of 235 participants. The mean age was 43.4 years (SD=13.0, range 14–71, see Figure 1A). 57.9% identified as male, 38.7% as female, 1.7% as diverse, and 1.7% selected the option stating that they did not wish to provide an answer (see Figure 1B). Participants were recruited by peers in open drug-use scenes (n=187, 79.6%), through posters in addiction clinics or substitution practices (n=42, 17.9%), or via unknown pathways (n=6, 2.6%). Respondents were located across large parts of Germany (see Figure 1C).

**Figure 1.**
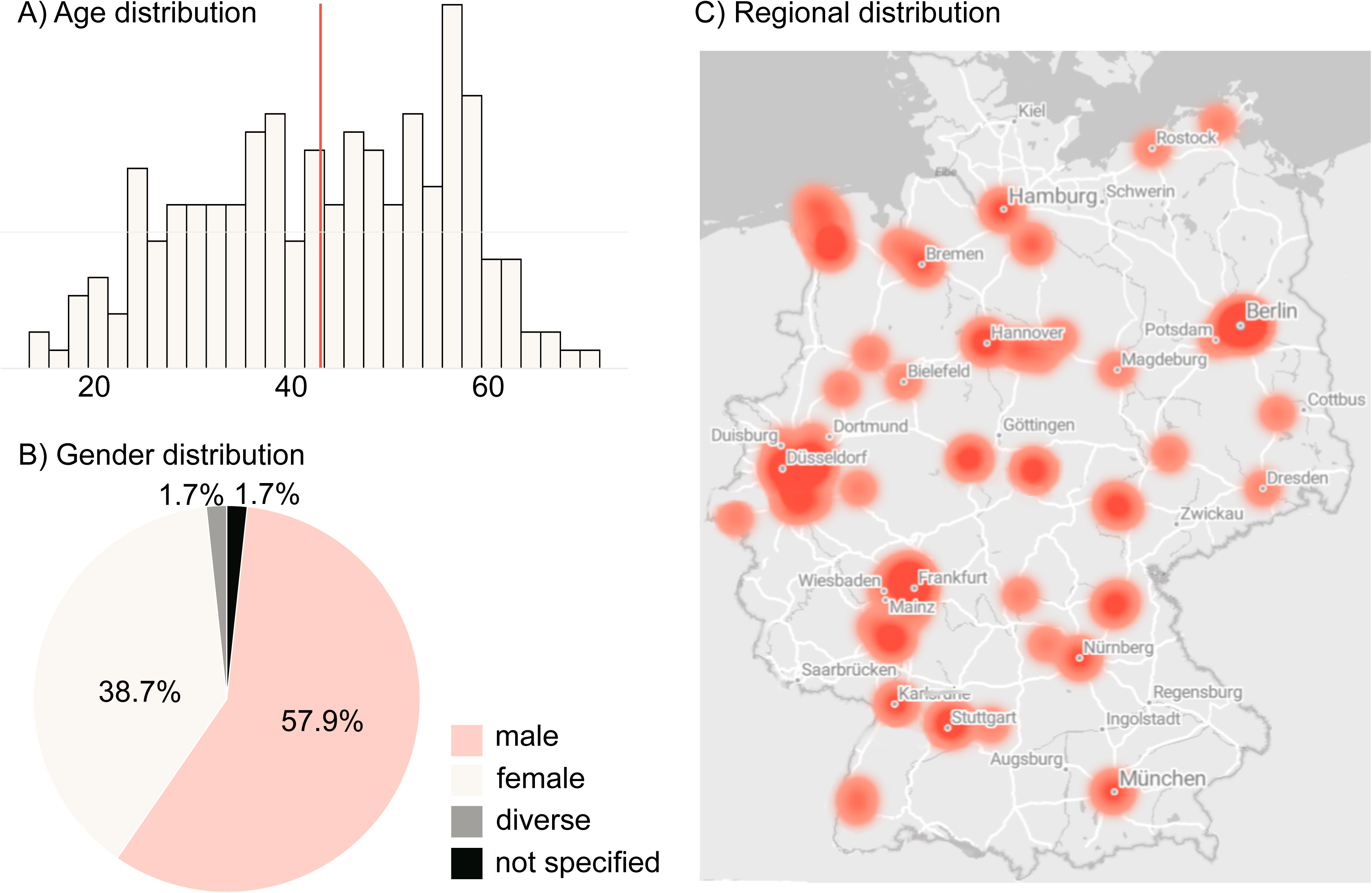
Sample characteristics. A) Frequency distribution of the respondents’ age. Each bar in the histogram represents two years of age; the red line indicates the mean age. B) Gender distribution in the sample. C) Regional distribution of respondents across Germany.

### Substance use in the past twelve months

Of the 235 participants, eight stated that they had not consumed any substances in the preceding twelve months and were therefore only shown questions not related to their own use. Among the other 227 participants, respondents reported using an average of 6.2 substances (SD=3.8; 5.8±3.5 for individuals recruited in the scene; 7.6± 4.3 for those from clinics/medical practices). After nicotine, alcohol, and cannabis, opioids for substitution (including methadone, levomethadone, buprenorphine, or diamorphine) and heroin were the most frequently mentioned substances (see Figure 2). Overall, 73.1% (95% CI=67.0–78.5%) reported consumption of at least one opioid. 11.0% used fentanyl or carfentanyl, and 4.8% nitazene-type opioids. Furthermore, the use of cocaine, sedatives, crack/freebase, stimulants, and pregabalin was widespread, with more than 30% of respondents reporting use of each substance (see Figure 2). For substances that can also be prescribed, the source of supply was inquired about. Apart from opioids for substitution, most of the substances were not obtained through medical care (see Supplementary Table S1).

**Figure 2.**
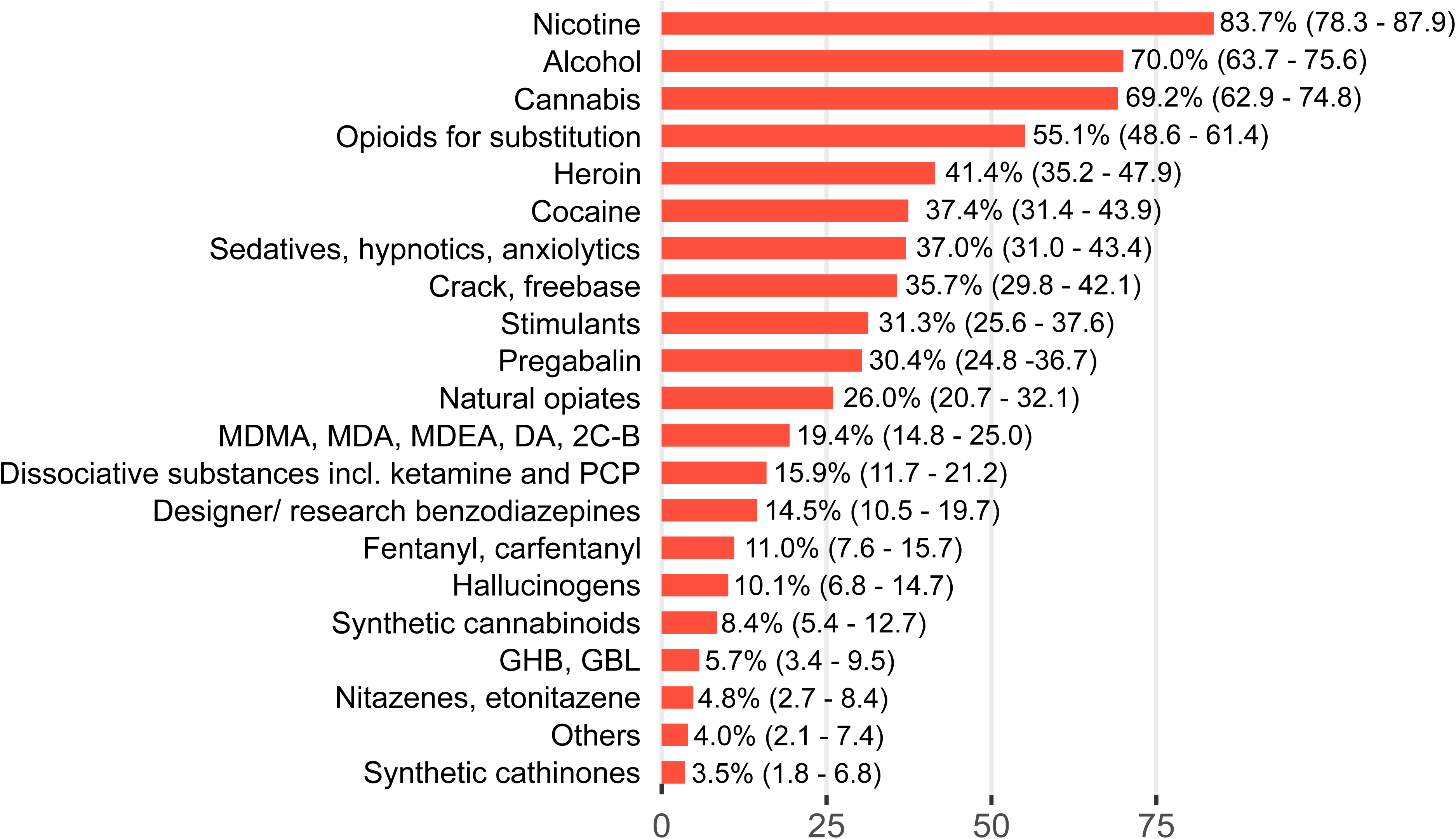
Percentages of respondents reported having used the respective substances in the past twelve months (95% Wilson confidence interval in parentheses).

Poly-substance use was also widespread among the 125 individuals taking opioids for substitution therapy with a mean of 4.0 additional substances (SD=3.5) excluding alcohol, nicotine, and cannabis. The most frequently co-used substances were heroin (53.6%, 95% CI=44.9–62.1%), sedatives (49.6%, 95% CI=41.0–58.2%), crack/freebase (46.4%, 95% CI=37.9–55.1%), and pregabalin (42.4%, 95% CI=34.1–51.2%).

### Substance use in the past twelve months stratified by age groups

The percentage of persons reporting consumption decreases with age for several substances, including alcohol, cannabis, cocaine, MDMA or related substances, dissociative substances, hallucinogens, and GHB/GBL (see Table 1). Teenagers and young adults have particularly high percentages for cocaine, stimulants, MDMA, hallucinogens, GHB/GBL, and synthetic cathinones compared to the other age groups. In contrast, they had the lowest proportion in the case of opioids for substitution, heroin, and crack/freebase. The consumption of sedatives, pregabalin, and designer/research benzodiazepines is highest in middle age, between 26 and 45 years. Synthetic opioids, both fentanyl/carfentanyl and nitazenes, are reported across all age groups (see Table 1).

**Table 1.** Percentage of individuals who reported consumption in the past 12 months, stratified by age group.

|  | age groups |  |  |  |  |
| --- | --- | --- | --- | --- | --- |
|  | <=25 y.<br>(n=25) | 26-35 y.<br>(n=41) | 36-45 y.<br>(n=54) | 46-55 y.<br>(n=56) | >55 y.<br>(n=51) |
| Nicotine | 80.0%<br>(60.9-91.1%) | 78.0%<br>(63.2-88.0%) | 90.7%<br>(80.0-96.0%) | 87.5%<br>(76.4-93.8%) | 78.4%<br>(65.3-87.5%) |
| Alcohol | 88.0%<br>(70.0-95.8%) | 75.6%<br>(60.6-86.2%) | 68.5%<br>(55.2-79.3%) | 73.2%<br>(60.4-83.0%) | 54.9%<br>(41.4-67.7%) |
| Cannabis | 84.0%<br>(65.3-93.6%) | 73.2%<br>(58.1-84.3%) | 70.4%<br>(57.2-80.9%) | 73.2%<br>(60.4-83.0%) | 52.9%<br>(39.5-65.9%) |
| Opioids for substitution | 20.0%<br>(8.9-39.1%) | 56.1%<br>(41.4-70.1%) | 59.3%<br>(46.0-71.4%) | 62.5%<br>(49.4-74.0%) | 58.8%<br>(45.1-71.2%) |
| Heroin | 20.0%<br>(8.9-39.1%) | 41.5%<br>(27.8-56.7%) | 50.0%<br>(37.1-62.9%) | 39.3%<br>(27.6-52.4%) | 45.1%<br>(32.3-58.6%) |
| Cocaine | 56.0%<br>(37.1-73.3%) | 41.5%<br>(27.8-56.7%) | 46.3%<br>(33.7-59.4%) | 33.9%<br>(22.9-47.0%) | 19.6%<br>(11.0-32.4%) |
| Sedatives, hypnotics,<br>anxiolytics | 36.0%<br>(20.2-55.5%) | 46.3%<br>(32.0-61.2%) | 51.9%<br>(38.9-64.7%) | 33.9%<br>(22.9-47.0%) | 17.6%<br>(9.5-30.2%) |
| Crack, freebase | 24.0%<br>(11.5-43.4%) | 46.3%<br>(32.0-61.2%) | 35.2%<br>(23.8-48.5%) | 41.1%<br>(29.2-54.2%) | 27.5%<br>(17.2-41.0%) |
| Stimulants | 56.0%<br>(37.1-73.3%) | 26.8%<br>(15.7-41.9%) | 50.0%<br>(37.1-62.9%) | 26.8%<br>(17.0-39.6%) | 7.8%<br>(3.1-18.4%) |
| Pregabalin | 24.0%<br>(11.5-43.4%) | 48.8%<br>(34.3-63.5%) | 46.3%<br>(33.7-59.4%) | 23.2%<br>(14.1-35.8%) | 9.8%<br>(4.3-21.0%) |
| Natural opiates | 32.0%<br>(17.2-51.6%) | 26.8%<br>(15.7-41.9%) | 33.3%<br>(22.2-46.6%) | 21.4%<br>(12.7-33.8%) | 19.6%<br>(11.0-32.4%) |
| MDMA, MDA, MDEA, DA,<br>2C-B | 40.0%<br>(23.4-59.3%) | 24.4%<br>(13.8-39.4%) | 27.8%<br>(17.6-40.9%) | 12.5%<br>(6.2-23.6%) | 3.9%<br>(1.1-13.2%) |
| Dissociative substances incl.<br>ketamine and PCP | 28.0%<br>(14.3-47.6%) | 26.8%<br>(15.7-41.9%) | 24.1%<br>(14.7-37.0%) | 5.4%<br>(1.9-14.7%) | 3.9%<br>(1.1-13.2%) |
| Designer/ research<br>benzodiazepines | 12.0%<br>(4.2-30.0%) | 17.1%<br>(8.5-31.3%) | 25.9%<br>(16.1-38.9%) | 10.7%<br>(5.0-21.5%) | 5.9%<br>(2.0-15.9%) |
| Fentanyl, carfentanyl | 12.0%<br>(4.2-30.0%) | 12.2%<br>(5.3-25.6%) | 14.8%<br>(7.7-26.6%) | 8.9%<br>(3.9-19.2%) | 7.8%<br>(3.1-18.4%) |
| Hallucinogens | 24.0%<br>(11.5-43.4%) | 9.8%<br>(3.9-22.6%) | 14.8%<br>(7.7-26.6%) | 7.1%<br>(2.8-16.9%) | 2.0%<br>(0.4-10.4%) |
| Synthetic cannabinoids | 12.0%<br>(4.2-30.0%) | 14.6%<br>(6.9-28.4%) | 13.0%<br>(6.4-24.5%) | 5.4%<br>(1.9-14.7%) | 0%<br>(0.0-7.0%) |
| GHB, GBL | 16.0%<br>(6.4-34.7%) | 12.2%<br>(5.3-25.6%) | 7.4%<br>(2.9-17.5%) | 0%<br>(0.0-6.4%) | 0%<br>(0.0-7.0%) |
| Nitazenes, etonitazene | 4.0%<br>(0.7-19.5%) | 4.9%<br>(1.4-16.2%) | 5.6%<br>(1.9-15.2%) | 7.1%<br>(2.8-16.9%) | 2.0%<br>(0.4-10.4%) |
| Others | 4.0%<br>(0.7-19.5%) | 4.9%<br>(1.4-16.2%) | 11.1%<br>(5.2-22.2%) | 0%<br>(0.0-6.4%) | 0%<br>(0.0-7.0%) |
| Synthetic cathinones | 8.0%<br>(2.2-25.0%) | 2.4%<br>(0.4-12.5%) | 3.7%<br>(1.0-12.5%) | 3.6%<br>(0.9-12.2%) | 2.0%<br>(0.4-10.4%) |

The values are percentages within each group, with 95% Wilson confidence intervals in parentheses.

### Substance use in the past twelve months stratified by gender

Substance use patterns were broadly similar across gender groups. With the exception of alcohol and pregabalin, where the proportion of female participants is about ten percentage points higher than that of male participants, slightly higher percentages were found among male respondents for most substances (see Table 2). Due to the very small sample size (n=4), no stratified analysis was conducted for individuals who identified as having a diverse gender.

**Table 2.** Percentage of individuals who reported consumption in the past 12 months, stratified by gender.

|  | gender |  |
| --- | --- | --- |
|  | male<br>(n=131) | female<br>(n=89) |
| Nicotine | 83.2%<br>(75.9-88.6%) | 86.5%<br>(77.9-92.1%) |
| Alcohol | 66.4%<br>(57.9-73.9%) | 76.4%<br>(66.6-84.0%) |
| Cannabis | 72.5%<br>(64.3-79.4%) | 64.0%<br>(53.6-73.2%) |
| Opioids for substitution | 58.8%<br>(50.2-66.9%) | 50.6%<br>(40.4-60.7%) |
| Heroin | 41.2%<br>(33.1-49.8%) | 41.6%<br>(31.9-52.0%) |
| Cocaine | 35.1%<br>(27.5-43.6%) | 40.4%<br>(30.8-50.8%) |
| Sedatives, hypnotics, anxiolytics | 38.9%<br>(31.0-47.5%) | 33.7%<br>(24.7-44.0%) |
| Crack, freebase | 36.6%<br>(28.8-45.1%) | 32.6%<br>(23.8-42.9%) |
| Stimulants | 35.9%<br>(28.2-44.4%) | 25.8%<br>(17.8-35.8%) |
| Pregabalin | 26.0%<br>(19.2-34.1%) | 36.0%<br>(26.8-46.4%) |
| Natural opiates | 25.2%<br>(18.5-33.3%) | 25.8%<br>(17.8-35.8%) |
| MDMA, MDA, MDEA, DA, 2C-B | 19.8%<br>(13.9-27.4%) | 18.0%<br>(11.4-27.3%) |
| Dissociative substances incl. ketamine and PCP | 16.0%<br>(10.7-23.2%) | 14.6%<br>(8.7-23.4%) |
| Designer/ research benzodiazepines | 14.5%<br>(9.5-21.5%) | 13.5%<br>(7.9-22.1%) |
| Fentanyl, carfentanyl | 11.5%<br>(7.1-18.1%) | 10.1%<br>(5.4-18.1%) |
| Hallucinogens | 13.7%<br>(8.8-20.6%) | 5.6%<br>(2.4-12.5%) |
| Synthetic cannabinoids | 10.7%<br>(6.5-17.2%) | 5.6%<br>(2.4-12.5%) |
| GHB, GBL | 6.1%<br>(3.1-11.6%) | 5.6%<br>(2.4-12.5%) |
| Nitazenes, etonitazene | 6.1%<br>(3.1-11.6%) | 2.2%<br>(0.6-7.8%) |
| Others | 3.1%<br>(1.2-7.7%) | 4.5%<br>(1.8-11.0%) |
| Synthetic cathinones | 6.1%<br>(3.1-11.6%) | 0%<br>(0.0-4.1%) |

The values are percentages within each group, with 95% Wilson confidence intervals in parentheses.

### Regional distribution of fentanyl and nitazenes use

Neither among individuals who reported using fentanyl/carfentanyl nor those using nitazenes was there a clustering in individual regions; rather a wide distribution across large parts of Germany was observed (see Figure 3).

**Figure 3.**
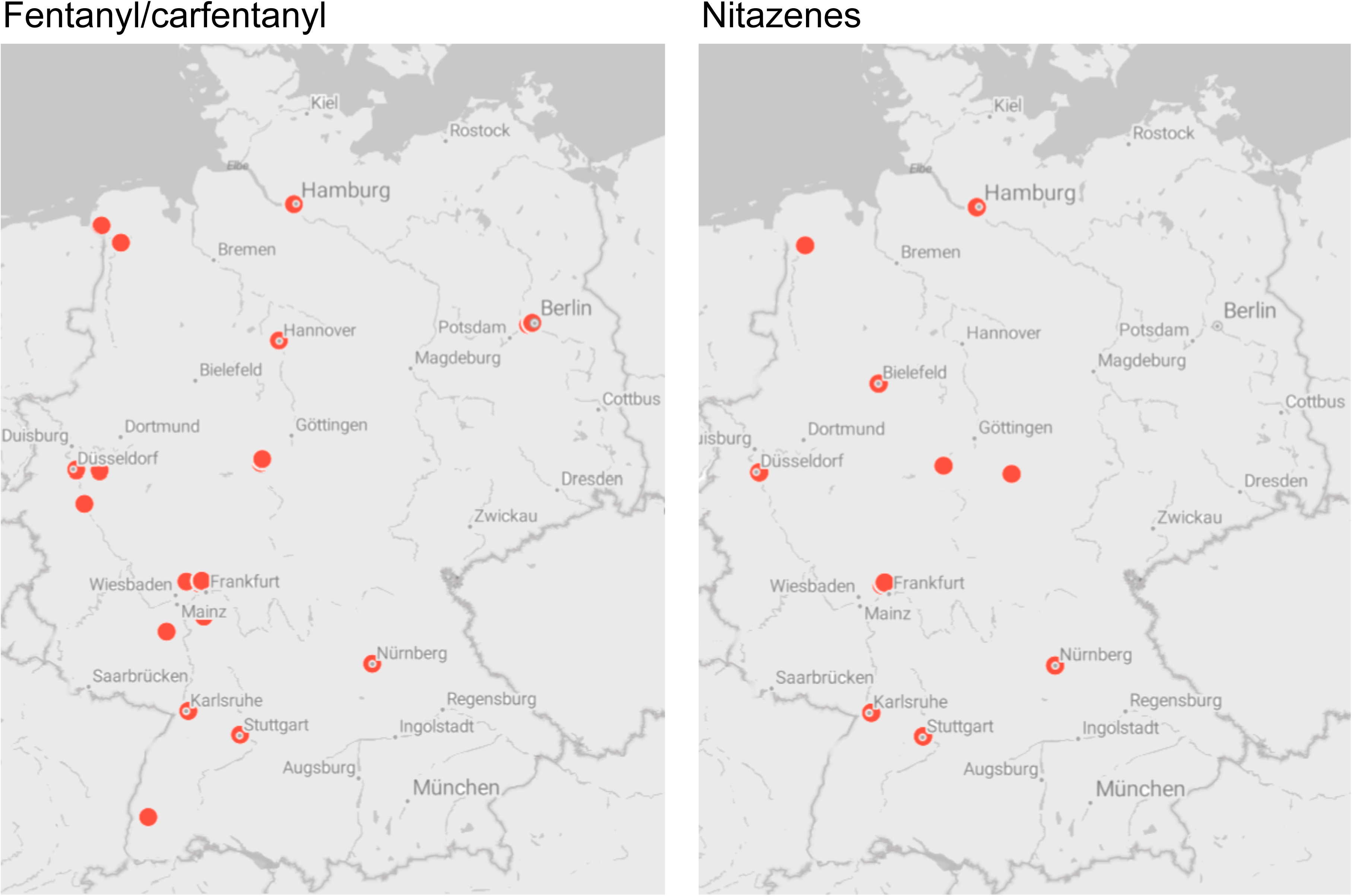
Regional distribution of individuals who reported using fentanyl/carfentanyl or nitazenes in the past 12 months.

Opioid use characteristics (opioid subgroup)

### Primary opioid and substitutes if the primary opioid was unavailable

Among the 166 participants who had consumed at least one opioid in the past 12 months, 159 provided information which opioid they considered their primary substance. The most frequently reported primary opioids were levomethadone, heroin, methadone, and diamorphine (see Table S2). Fentanyl was chosen by 1.9%, nitazenes by 1.3%. When asked whether it had happened that they consumed another substance because the primary substance was unavailable, 38.2% (95% CI=31.0– 46.0%) answered yes (of n=157 valid responses). Among these individuals (n=60), heroin, methadone, levomethadone, and fentanyl were most often selected as substitute (see Table S3). Among respondents who also selected sources other than medical care, 41.5% (95% CI=30.3-53.6%) reported decreased availability of their primary opioid (n=65). Detailed information about supply sources, form, route of use, perceived availability, and price development of the primary opioid can be found in Table S4.

### Fentanyl/carfentanyl and nitazenes sources, form, route of use, and availability (subgroup who had consumed the respective substance)

Of those individuals who had reported fentanyl/carfentanyl consumption in the past 12 months (n=25), 80.0% indicated the illicit market/street as source of supply (see Table S5). Among those who did not report medical care as their sole source of supply, the most common methods of use were inhalation and injection. 54.5% indicated that the availability of fentanyl/carfentanyl had increased (see Figure 4A and S5). Of those who had consumed nitazenes, 4 of 9 valid cases (44.4%, 95% CI=18.8-73.3%) stated that the availability had increased (see Figure 4A and Table S5). Nitazenes were most commonly consumed by injection and sniffing. For more information about supply source, form, route of use, perceived availability, and price development of fentanyl/carfentanyl and nitazenes see Table S5.

**Figure 4.**
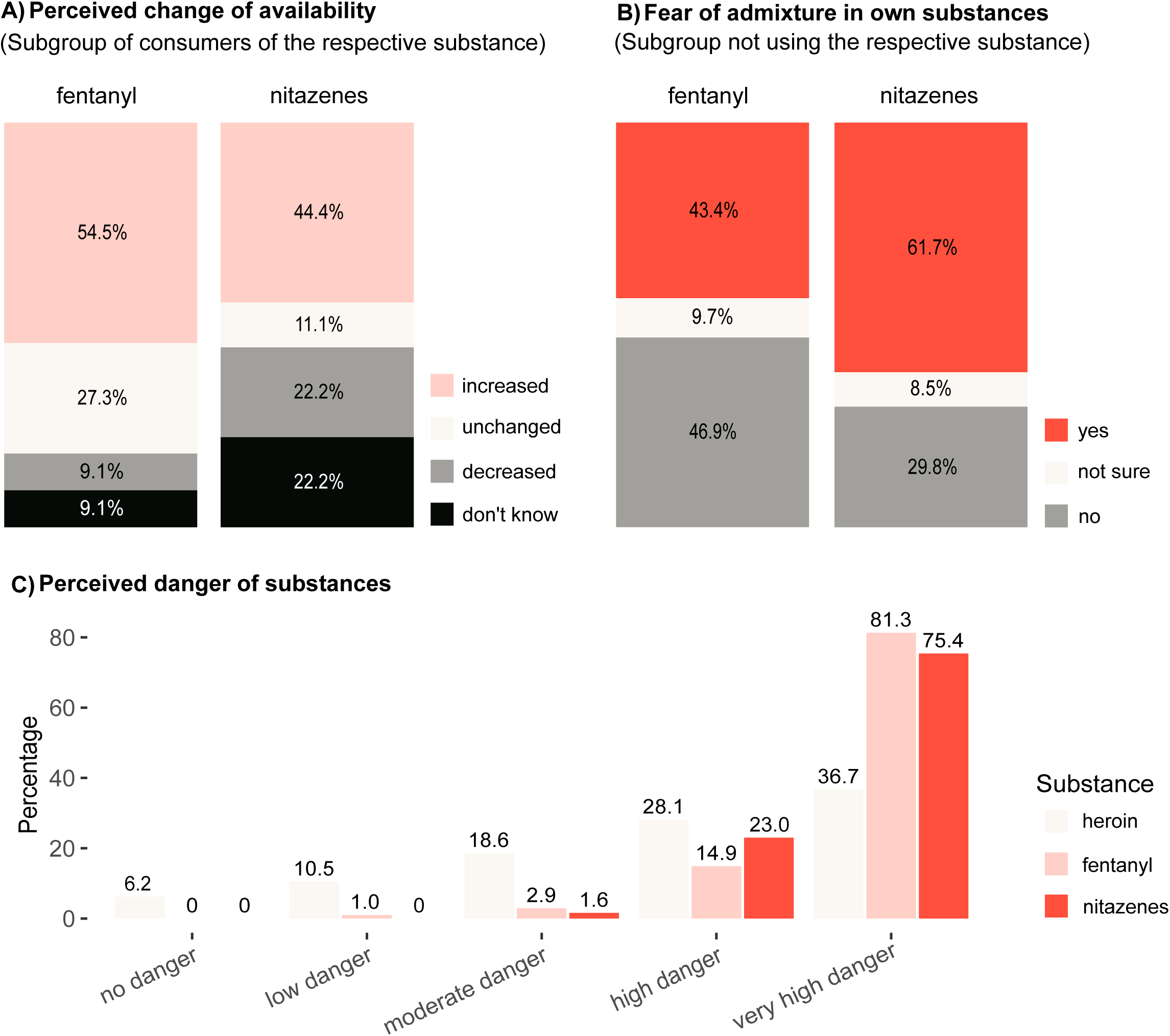
Perceived availability and awareness of risks associated with fentanyl and nitazenes. A) Changes in availability as reported by consumers of the respective substance. Individuals who obtained fentanyl exclusively through medical care were not part of this analysis, resulting in sample sizes of n=22 for fentanyl and n=9 valid cases for nitazenes. B) Percentage of those who are afraid of adulteration among individuals who have not used the respective substance in the past twelve months but are familiar with it (n=175 for fentanyl, n=47 for nitazenes). C) Ratings indicating how dangerous the respective substance is considered to be among both individuals using it and persons who don’t use the substance but are familiar with it (n=210 for heroin, 208 for fentanyl, and 61 for nitazenes).

Fentanyl- and nitazene-related awareness and perceived availability (subgroup who had not consumed the respective substance)

Among respondents who had not consumed fentanyl in the past twelve months, 91.2% of 194 valid cases reported knowing the substance (see S6). Of these, 54.0% stated that fentanyl is available in their region (see S6) and a substantial proportion feared that fentanyl could be mixed into their own substances (see Figure 4B and S6). Of respondents who had not taken nitazenes in the past twelve months, 66.8% of 208 valid cases did not know these substances (see S6). Among those familiar with nitazenes (n=49), many were uncertain about regional availability (44.9%), but 40.8% confirmed it (see S6) and the majority feared adulteration (see Figure 4B and S6). More information about awareness, interest, and perceived availability of fentanyl and nitazenes can be found in S6.

### Risk perception, risk behaviors and harm-reduction indicators

Regardless of whether they had already used the substances, participants were asked how dangerous they considered heroin, fentanyl, and nitazenes. Synthetic opioids were more frequently rated as very high danger than heroin (see Figure 4C and Table S7). With regard to different forms of consumption, intravenous consumption was rated as high or very high risk by the vast majority of participants, while for inhalation, nasal, and oral consumption, a medium risk was most frequently indicated (see S7). 55.5% (95% CI=48.7–62.1%) of the respondents (n=209 valid cases) reported that they mostly consume alone and 19.7% stated that they currently use drug consumption rooms (see Figure 5); of opioid users it was 25.2% (of n=151 valid cases, 95% CI= 18.9-32.7%). Among those who do not use consumption rooms, 35.3% stated that they would use them if they were available in their region (see Figure 5); among opioid users it was 43.4% (of 113 valid cases, 95% CI=34.6-52.6%). 9.9% of all respondents use drug checking, but 75.7% of those who said they do not would use the service if it were available in their region (see Figure 5). When asked whether they owned a naloxone emergency kit, 30.9% (95% CI=24.1–38.6%) of opioid users (n=152 valid cases) answered in the affirmative.

**Figure 5.**
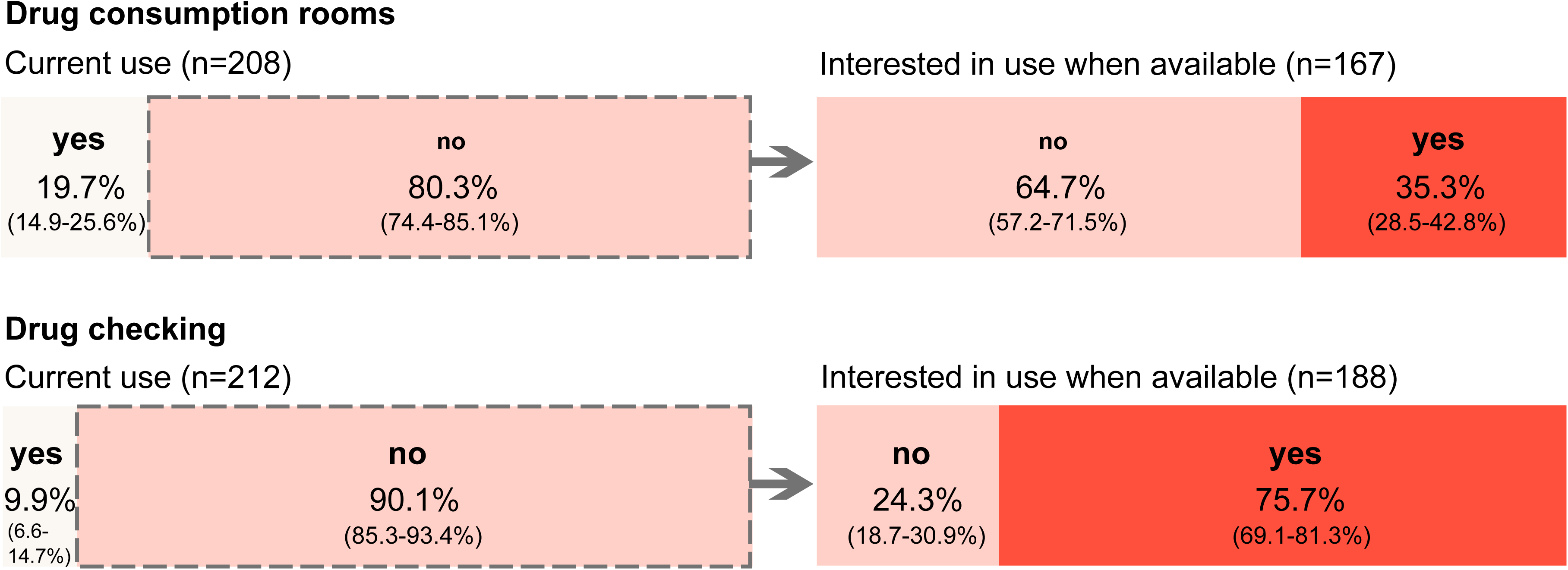
Current use of and interest in drug consumption rooms and drug checking. All individuals who reported not using these services were asked whether they would use them if they were available in their region.

Overdose experiences, overdose concerns, and suicide attempts 80.5% (95% CI=74.6–85.3%) of respondents had heard about overdoses in others and 40.0% (95% CI=33.6–46.7%) had witnessed at least one in the past twelve months (n=210 valid responses). 67.9% (95% CI=61.3-73.9%) reported concerns about an overdose in themselves or others and 60.6% (95% CI=52.4-68.3%) of those concerned (n=142) stated they were more concerned than they used to be. Personal overdose in the past 12 months was reported by 16.3% (95% CI=11.8-22.0%) of respondents (n=202 valid responses); among users of synthetic opioids, the proportion was 56.5% (95% CI=36.8-74.3%). Of all respondents (n=194 valid cases), 9.8% (95% CI=6.4-14.8%) reported having attempted suicide in the past twelve months. Among users of synthetic opioids, this figure was 22.7% (95% CI=10.1-43.4%, n=22). Seven of eleven users of synthetic opioids who reported an overdose in the past twelve months stated they had not attempted suicide.

## DISCUSSION

The present study reports findings from a cross-sectional survey conducted at multiple locations within Germany’s drug-use scene. Our study found extensive polysubstance use, with participants reporting a mean of more than six substances. Regarding synthetic opioids, 11.0% of respondents reported use of fentanyl or carfentanyl and 4.8% reported use of nitazene-type opioids in the past 12 months. Among those who used synthetic opioids, more than half reported having experienced an overdose in the past 12 months.

Our data on fentanyl/carfentanyl use indicate that consumption is already widespread across large parts of Germany and across all age and gender groups, with 80% reporting the illicit market as their source of supply. The prevalence of synthetic opioid use in our sample remains far below that reported from North America, consistent with monitoring data [16] showing that heroin is still the most commonly used illicit opioid and that crack is also highly prevalent. Several factors likely contribute to this divergence from North American patterns, including longstanding heroin trafficking routes from Afghanistan via Turkey and the Balkans [17–19]. At the same time, it is concerning that most fentanyl users in this survey reported increasing availability. Combined with recent evidence that fentanyl is already present as an adulterant in tested heroin samples in Germany, this raises concern that exposure may rise in the coming years [20]. Notably, a considerable proportion of respondents who obtain their primary opioid from the illicit market reported declining availability, and among those who reported a shortage of their primary opioid in the past year, one quarter stated they had used fentanyl instead - both of which are trends that were also identified in a recent survey among experts [21]. Given its high potency, such market-driven switching may substantially increase overdose risk and is highly relevant to ongoing debates on how supply disruptions, such as those following the Taliban’s ban on opium poppy cultivation, may shape opioid-related harms in Europe [22].

Nitazenes have been highlighted as a particular concern for Europe because of their extreme potency [23]. In our sample, nearly one in twenty respondents reported nitazene use in the past 12 months, with all age and gender groups and many parts of Germany represented. However, they remain far less familiar than fentanyl, with about two-thirds of respondents unaware of them. Among those who were familiar with nitazenes, perceived risk was high and concern about adulteration was widespread. This indicates that, while serious concerns exist within parts of the community, low awareness among PWUD may impair the ability to recognize risks associated with these highly potent opioids and to respond appropriately.

A key finding of this survey is the high prevalence of polysubstance use, including frequent concurrent use of sedatives/anxiolytics, pregabalin, and stimulants. The combination of opioids with other respiratory depressants can markedly increase overdose risk [24–27], and stimulant-opioid combinations may further complicate toxicity profiles and risk behaviors [28–30]. Polysubstance use was also widespread among individuals in OAT, with an average of four concurrent substances (excluding alcohol, nicotine and cannabis). These findings suggest that current treatment settings may not yet adequately address polysubstance use and underscore the need for stronger routines for identifying and addressing concurrent use of benzodiazepines, alcohol, pregabalin, and stimulants.

The present data indicate that risky patterns of use are common, while harm-reduction measures are rarely used. More than one third of opioid users reported using their primary opioid intravenously, more than half of all respondents stated mostly consuming alone and the minority reported using drug consumption rooms. Drug checking was used by only 9.9%, and less than one third of opioid users owned a naloxone emergency kit. The danger of this pattern is underscored by overdose rates. 16.3% of all respondents reported having experienced an overdose in the past 12 months, among synthetic opioid users, this proportion rose to 56.5%. Of those who reported an overdose, approximately two-thirds reported not having attempted suicide in the preceding 12 months, suggesting that the majority of overdoses involving synthetic opioids were unintentional. These findings are consistent with international mortality analyses. Moreover, a large proportion of respondents reported being more concerned about overdose, for themselves or others, than they used to be.

Taken together, these findings clearly indicate a need for action. First, drug checking should be expanded, as it reduces overdose risk and other drug-related harms [31–33]; however, it is currently available only on a limited basis in Germany and is not regulated by law in all federal states. Three quarters of those who do not currently use drug checking stated that they would do so if it were available in their region, indicating a clear unmet demand. Second, naloxone distribution and related training should be expanded, particularly in settings characterized by polysubstance use and high uncertainty regarding opioid content, including low threshold drug services, OAT programmes, emergency shelters, prisons, probation services, hospitals, and outpatient practices [11,28,34]. Third, training for emergency physicians and other frontline staff (e.g. ambulance services, emergency departments, police, and street-based services) is needed to improve recognition and management of overdoses involving highly potent synthetic opioids. Forth, regional availability of drug consumption rooms should be supported and access barriers reduced as nearly 20% of respondents already use these facilities and more than one third of non-users would do so if they were available for them. Fifth, polysubstance use should be addressed systematically, particularly within OAT, through combined pharmacological and psychosocial interventions. Sixth, early warning systems for synthetic opioids should be reinforced through enhanced toxicological screening and rapid dissemination of alerts to local services and communities, complemented by expanded targeted information for PWUD about substances, risks, safer use strategies, and appropriate overdose responses. These recommendations are broadly aligned with the United Nations synthetic drug strategy [35], which emphasizes multiindicator early warning, strengthened health responses, and enhanced capacities to detect and respond to highly potent synthetic substances.

This study has several strengths. Recruitment through peers and low-threshold services enabled access to individuals who are often insufficiently represented in general population surveys. The survey was developed with input from an interdisciplinary expert panel and people with lived experience, supporting content validity and acceptability. At the same time, several limitations must be considered when interpreting the results. The sampling strategy was non-probabilistic, which limits representativeness and may have introduced selection bias toward particular networks or service-connected groups. In addition, all measures were self-reported, raising the possibility of recall error, stigma-related underreporting, and misclassification [36]. For example, fentanyl or nitazene exposure may be suspected but not analytically confirmed or may have occurred unknowingly through adulteration.

## Conclusion

In conclusion, this scene-based survey provides evidence that fentanyl/carfentanyl and nitazenes are already present across multiple drugUuse scenes in Germany and are accompanied by high rates of non-fatal overdose, substantial concern among PWUD, and insufficient availability of harm-reduction services. Aligning with European and international guidance on synthetic drugs, our findings underscore the need to strengthen multi-indicator monitoring and to expand integrated harm-reduction and treatment responses to mitigate the risk of an escalation of opioid-related harms in Germany.

## Data Availability

All data produced in the present study are available upon reasonable request to the authors.

## ACKNOWLEDGMENTS

We would like to express our special thanks to all our partners, especially

- all voluntary participants, peers and helpers for answering the questions, their support with recruitment and the conduct of the interviews, in particular Nils Badenheuer, Roland Baur, Heike Bembennek, Jennifer Blaine, Timo Eichel, and Martina Tranel;
- all members of the JES-Bundesverband e.V., especially Claudia Ak;
- all members of the Deutsche Aidshilfe, especially Maria Kuban and Dirk Schäffer;
- Hilfe zur Selbsthilfe – Begegnung Jena e.V.;
- Sozialdienst katholischer Frauen und Männer Düsseldorf e.V.;
- all participating addiction clinics and specialized practices, in particular Schwerpunktpraxis für Suchtmedizin Stuttgart, Dr. Andreas Zsolnai;
- the expert panel for their support in developing the questionnaire and their helpful advice regarding the implementation of the survey, in particular Andreas Heinz, Michael Krausz, Diana Moesgen, Lenea Reuvers, Norbert Scherbaum, Katharina Schoett, and Sven Speerforck.

## STATEMENT OF ETHICS

The study protocol was approved by the Ethics Committee of the State Medical Association (Landesärztekammer) Baden-Württemberg (approval number: F-2024-120).

## AUTHORS’ CONTRIBUTIONS

Conceptualization: JR, LS, NW, LR, MC. Methodology: JR, LS, NW, LR, MC. Investigation: NW, LR, LS, JR.

Data curation: LS, LR, NW. Formal analysis: LS, LR.

Project administration: JR, LR, LS. Writing – original draft: JR, LR, LS.

Writing – review & editing: JR, LR, LS, MC.

All authors contributed to the development of the questionnaire in collaboration with the expert group and approved the final version of the manuscript.

## DATA AVAILABILITY STATEMENT

The data that support the findings of this study are not publicly available due to privacy reasons but are available from the evaluation center upon request. Further enquiries can be directed to the corresponding author.

