## Supplemental material for "Opioid crisis in Germany? Insights from a cross-sectional nationwide survey within the German drug scene"

Supplementary material

S1. Source of consumed substances that can be prescribed.

|  | prescribed | scene/ street | internet | not specified |
| --- | --- | --- | --- | --- |
| Cannabis (n=157) | 10.8%  (6.8-16.6%) | 61.1%  (53.3-68.4%) | 5.1%  (2.6-9.7%) | 31.2%  (24.5-38.8%) |
| Opioids for substitution (n=125) | 66.4%  (57.7-74.1%) | 13.6%  (8.7-20.7%) | 0.8%  (0.1-4.4%) | 24.8%  (18.1-33.0%) |
| Sedatives, hypnotics, anxiolytics (n=84) | 34.5%  (25.2-45.1%) | 46.4%  (36.1-57.0%) | 3.6%  (1.2-10.0%) | 28.6%  (20.0-39.0%) |
| Stimulants (n=71) | 7.0%  (3.0-15.4%) | 67.6%  (56.1-77.3%) | 2.8%  (0.8-9.7%) | 29.6%  (20.3-41.0%) |
| Pregabalin (n=69) | 15.9%  (9.1-26.3%) | 63.8%  (52.0-74.1%) | 1.4%  (0.2-7.7%) | 24.6%  (16.0-35.9%) |
| Natural opiates (n=59) | 30.5%  (20.2-43.1%) | 42.4%  (30.6-55.1%) | 1.7%  (0.3-9.0%) | 33.9%  (23.1-46.6%) |
| Fentanyl, cerfentanyl (n=25) | 4.0%  (0.7-19.5%) | 68.0%  (48.4-82.8%) | 4.0%  (0.7-19.5%) | 28.0%  (14.3-47.6%) |

It was possible to select multiple sources for each substance. The “not specified” column shows the percentage of people who did not answer the question.

S2. Opioid rated as primary by opioid users (n=159).

| Levomethadone | 23.3% (17.4-30.5%) |
| --- | --- |
| Heroin | 18.2% (13.0-24.9%) |
| Methadone | 13.2% (8.8-19.3%) |
| Diamorphine | 13.2% (8.8-19.3%) |
| Buprenorphine | 7.5% (4.3-12.7%) |
| Morphine | 7.5% (4.3-12.7%) |
| Others | 6.3% (3.5-11.2%) |
| Tilidin | 2.5% (1.0-6.3%) |
| Codein | 1.9% (0.7-5.4%) |
| Fentanyl | 1.9% (0.7-5.4%) |
| Tramadol | 1.9% (0.7-5.4%) |
| Nitazenes | 1.3% (0.4-4.5%) |
| Oxycodon | 1.3% (0.4-4.5%) |

Percentages with 95% Wilson confidence intervals in parentheses.

S3. Opioids consumed as substitutes when the primary substance was not available (n=60, subgroup of person who reported it had happened that their primary substance was unavailable in the past twelve months).

| Heroin | 41.7% (30.1-54.3%) |
| --- | --- |
| Methadone | 31.7% (21.3-44.3%) |
| Levomethadone | 26.7% (17.2-39.0%) |
| Fentanyl | 25.0% (15.8-37.2%) |
| Oxycodon | 21.7% (13.1-33.7%) |
| Tilidin | 21.7% (13.1-33.7%) |
| Buprenorphine | 16.7% (9.3-28.1%) |
| Other* | 15.0% (8.1-26.1%) |
| Tramadol | 10.0% (4.7-20.1%) |
| Diamorphine | 6.7% (2.6-16.0%) |
| Nitazenes | 6.7% (2.6-16.0%) |
| Morphine | 6.7% (2.6-16.0%) |
| Opium | 6.7% (2.6-16.0%) |
| Carfentanyl | 3.3% (0.9-11.3%) |
| Codein | 3.3% (0.9-11.3%) |

Percentages with 95% Wilson confidence intervals in parentheses. Multiple responses were possible; up to nine alternatives were selected.

*cannabis, crack , benzodiazepines, and pregabalin were mentioned as other substances

S4. Supply source, form, route of use, perceived availability, and price development of primary opioid

| Source of supply (n=154) * | Medical care | 63.9% (56.1-71.1%) |
| --- | --- | --- |
|  | Drug scene/street | 34.4% (27.4-42.2%) |
|  | Family/friends | 13.6% (9.1-19.9%) |
|  | Internet | 1.3% (0.4-4.6%) |
|  | Other | 2.6% (1.0-6.5%) |
| Form (n=64)* | Powder | 53.1% (41.0-64.8%) |
|  | Tablets | 26.6% (17.3-38.5%) |
|  | Drops | 20.3% (12.3-31.7%) |
|  | Patches | 6.3% (2.5-15.1%) |
|  | Other | 9.4% (4.4-19.0%) |
| Route of use (n=64)* | Intravenous consumption | 35.9% (25.3-48.1%) |
|  | Swallowing | 34.4% (23.9-46.6%) |
|  | Inhaling | 26.6% (17.3-38.5%) |
|  | Sniffing | 23.4% (14.7-35.1%) |
|  | Sucking | 4.7% (1.6-12.9%) |
|  | Sticking | 1.6% (0.3-8.4%) |
|  | Boiling before consumption | 9.4% (4.4-19.0%) |
| Perceived change of availability (n=65) | Increased | 10.8% (5.3-20.6%) |
|  | Unchanged | 36.9% (26.2-49.1%) |
|  | Decreased | 41.5% (30.3-53.6%) |
|  | Not available anymore | 1.5% (0.3-8.2%) |
|  | Don’t know | 9.2% (4.3-18.7%) |
| Perceived price change (n=65) | Increased | 23.1% (14.5-34.7%) |
|  | Unchanged | 46.2% (34.6-58.2%) |
|  | Decreased | 15.4% (8.6-16.1%) |
|  | Don’t know | 15.4% (8.6-16.1%) |

Percentages with 95% Wilson confidence intervals in parentheses.

*multiple selections possible

n=88 (57.1%) reported medical care as their sole source of supply and were therefore not asked the following questions about forms of consumption and availability/price. Two persons did not choose any option for form and route of rouse and one person for availability and price change, resulting in sample sizes of 64 and 65, respectively.

S5. Supply source, form, route of use, perceived availability, and price development of fentanyl and nitazenes

|  |  | Fentanyl | Nitazenes |
| --- | --- | --- | --- |
| Source of supply * | Medical care | 16.0% (6.4-34.7%) | - |
|  | Drug scene/street | 80.0% (60.9-91.1%) | 75.0% (40.9-92.9%) |
|  | Family/friends | 12.0% (4.2-30.0%) | 25.0% (7.15-59.1%) |
|  | Internet | 4.0% (0.7-19.5%) | 50.0% (21.5-78.5%) |
|  | Other | 4.0% (0.7-19.5%) | 12.5% (2.2-47.1%) |
| Form* | Powder | 28.6% (13.8-50.0%) | 87.5% (52.9-97.8%) |
|  | Tablets | 0% (0.0-15.5%) | 12.5% (2.2-47.1%) |
|  | Drops | 4.8% (0.9-22.7%) | 37.5% (13.7-69.4%) |
|  | Patches | 90.5% (71.1-97.4%) | 12.5%(2.2-47.1%) |
|  | Other | 0% (0.0-15.5%) | 0% (0.0-32.4%) |
| Route of use* | Intravenous consumption | 47.6% (28.3-67.6%) | 57.1% (25.0-84.2%) |
|  | Swallowing | 4.8% (0.9-22.7%) | 28.6% (8.2-64.1%) |
|  | Inhaling | 47.6% (28.3-67.6%) | 42.9% (15.8-75.0%) |
|  | Sniffing | 4.8% (0.9-22.7%) | 57.1% (25.0-84.2%) |
|  | Sucking | 38.1% (20.8-59.1%) | - |
|  | Sticking | 14.3% (4.9-34.5%) | 14.3% (2.6--51.3%) |
|  | Boiling before consumption | 33.3% (17.2-54.6%) | - |
| Perceived change of availability | Increased | 54.5% (34.6-73.0%) | 44.4% (18.8-73.3%) |
|  | Unchanged | 27.3% (13.2-48.2%) | 11.1% (2.0-43.5%) |
|  | Decreased | 9.1% (2.5-27.8%) | 22.2% (6.3-54.7%) |
|  | Not available anymore | 0% (0.0-14.9%) | 0% (0.0-29.9%) |
|  | Don’t know | 9.1% (2.5-27.8%) | 22.2% (6.3-54.7%) |
| Perceived price change | Increased | 22.7% (10.1-43.4%) | 0% (0.0-29.9%) |
|  | Unchanged | 40.9% (23.2-61.3%) | 33.3% (12.0-64.6%) |
|  | Decreased | 4.5% (0.8-21.7%) | 11.1% (2.0-43.5%) |
|  | Don’t know | 31.8% (16.3-52.7%) | 55.6% (26.7-81.2%) |

Percentages with 95% Wilson confidence intervals in parentheses. *multiple selections possible

n=25 for source of fentanyl. Three individuals stated that medical care was their sole source of supply and were not asked the other questions. Furthermore, one subject did not select any response option for form and route of administration, resulting in n=21 for these two questions and n=22 for perceived change of availability and price of fentanyl. Of those who had consumed nitazenes (n=11), n=8 answered the questions about source and form, n=7 the question about route of admininstration, and n=9 those about peceived change of availability and price of nitazenes.

S6 Fentanyl- and nitazene-related awareness, interest, and perceived availability (subgroup who had not consumed the respective substance)

Participants who had not consumed fentanyl in the past twelve months were asked whether they knew what fentanyl was. Of n=194 respondents, 91.2% (95% CI=86.4-94.4%; n=177) answered yes, 6.7% (95% CI = 4.0-11.1%) no, and 2.1% (95%CI= 0.8-5.2%) were unsure. Among those who were familiar with fentanyl, 45.7% (95% CI=38.5-53.1%) stated that they discussed fentanyl among their peers, while 49.7% (95% CI=42.4-57.0%) reported no discussion and 4.6% (95% CI=2.4-8.8%) were unsure (n=175 valid cases). 12.5% (95% CI=8.4-18.2%) responded that they were interested in trying fentanyl (again, if applicable), whereas 80.7% (95% CI=74.2-85.8%) expressed no interest, and 6.8% (95% CI=3.9-11.5%) were unsure (n=176 valid cases). When asked whether fentanyl is available in their region, 54.0% (95% CI=46.6-61.2%) answered yes, 10.2% (95% CI=6.5-15.6%) answered no, and 35.8% (95% C= 29.1-43.1%) were unsure (n=176 valid cases). 43.4% (95% CI=36.3-50.8%) expressed fear that fentanyl could be mixed into their own substances, 46.9% had no fear (95% CI=39.7-54.3%) and 9.7% were not sure (95% CI=6.1-15.0%).

All individuals who had not taken nitazenes in the past twelve months were also asked if they knew what nitazenes were. Of 208 valid cases, 66.8% (95% CI=60.1-72.8%) replied “no” to this question, 23.6% (95% CI=18.3-29.8) replied “yes”, and 9.6% (95% CI=6.3-14.4%) were unsure. Of those who were familiar with nitazenes, 62.5% (95% CI=48.4-74.8%) stated that they discussed them with their peers, 35.4% (95% CI=23.4-49.5%) indicated that they did not, and 2.1% (95% CI=0.4-10.9%) were unsure (n=48 valid cases). Asked whether they were interested in trying nitazenes (again, if applicable), 81.3% (95% CI=68.1-89.8%) indicated that they were not interested, 10.4% (95% CI=4.5-22.1%) expressed interest, and 8.3% (95% CI=3.3-19.5%) were unsure (n=48 valid cases). The largest proportion of them (44.9%, 95% CI=31.9-58.7%) did not know whether nitazenes were available in their region; 40.8% (95% CI=28.2-54.7%) confirmed the availability, and 14.3% (95% CI=7.1-26.7%) denied this (n=49). 61.7% (95% CI=47.4-74.2%) feared that nitazenes could be mixed into their own substances, 29.8% had no fear (95% CI=18.7-44.0%) and 8.5% were not sure (95% CI=3.4-19.9%).

S7 Perceived danger of different routes of application and of synthetic opioids compared to heroin

|  | no danger | low danger | moderate danger | high danger | very high danger |
| --- | --- | --- | --- | --- | --- |
| Intravenous use (n=210) | 1.0%  (0.3-3.5%) | 0%  (0.0-1.8%) | 8.1%  (5.1-12.6%) | 20.5%  (15.6-26.5%) | 70.5%  (64.0-76.3%) |
| Inhalation (n=210) | 2.4%  (1.0-5.5%) | 18.1%  (13.5-23.9%) | 37.6%  (31.3-44.3%) | 24.8%  (19.4-31.1%) | 17.1%  (12.6-22.8%) |
| Nasal use (n=209) | 2.4%  (1.0-5.5%) | 16.7%  (12.3-22.3%) | 36.8%  (30.6-43.5%) | 24.4%  (19.1-30.7%) | 19.6%  (14.8-25.5%) |
| Oral use (n=207) | 5.8%  (3.3-9.9%) | 18.8%  (14.1-24.7%) | 37.2%  (30.9-44.0%) | 18.8%  (14.1-24.7%) | 19.3%  (14.5-25.2%) |
| Heroin (n=210) | 6.2%  (3.7-10.3%) | 10.5%  (7.0-15.4%) | 18.6%  (13.9-24.4%) | 28.1%  (22.5-34.5%) | 36.7%  (30.5-43.4%) |
| Fentanyl (n=208) | 0%  (0.0-1.8%) | 1.0%  (0.3-3.5%) | 2.9%  (1.3-6.2%) | 14.9%  (10.7-20.4%) | 81.3%  (75.5-86.0%) |
| Nitazenes (n=61) | 0%  (0.0-5.9%) | 0%  (0.0-5.9%) | 1.6%  (0.3-8.7%) | 23.0%  (14.2-35.0%) | 75.4%  (63.3-84.5%) |

Percentages with 95% Wilson confidence intervals in parentheses.
